# The Proteome Landscape of Human Placentas for Monochorionic Twins with Selective Intrauterine Growth Restriction

**DOI:** 10.1101/2022.08.29.22278892

**Authors:** Xin-Lu Meng, Peng-Bo Yuan, Xue-Ju Wang, Jing Hang, Xiao-Ming Shi, Yang-Yu Zhao, Yuan Wei

## Abstract

In perinatal medicine, intrauterine growth restriction (IUGR) is one of the greatest challenges. The etiology of IUGR is multifactorial, but most cases are thought to arise from placental insufficiency. However, identifying the placental cause of IUGR can be difficult due to numerous confounding factors. Selective IUGR (sIUGR) would be a good model to investigate how impaired placentation affects fetal development, as the growth discordance between monochorionic twins cannot be explained by confounding genetic or maternal factors. Herein we constructed and analyzed the placental proteomic profiles of IUGR twins and the normal cotwins. Specifically, we identified a total of 5481 proteins and 233 differentially expressed proteins (DEPs), including 57 upregulated and 176 downregulated DEPs in IUGR twins. Bioinformatic analysis indicates that these DEPs are mainly associated with cardiovascular system development and function, organismal survival, and organismal development. Notably, 34 DEPs are significantly enriched in angiogenesis, and diminished placental angiogenesis in IUGR twins has been further elaborately confirmed. Moreover, we found decreased expression of *metadherin* (*MTDH*) in placentas for IUGR twins and demonstrated that *MTDH* contributes to placental angiogenesis and fetal growth *in vitro*. Collectively, our findings reveal the comprehensive proteomic signature of placentas for sIUGR twins, and the DEPs identified may provide in-depth insights into pathogenesis of placental dysfunction and subsequent impaired fetal growth.

## Introduction

Intrauterine growth restriction (IUGR), which refers to the inability of the fetus to reach its full growth potential, is among the most common pregnancy complications and affects 7% of pregnancies worldwide [1]. Besides contributing to significant perinatal and pediatric morbidity and mortality, it can also raise the risk of developing metabolic, cardiovascular, and respiratory disorders later in life [2,3]. Despite the involvement of multiple factors, placental insufficiency has been reported as a major cause of IUGR [4,5]. Placental insufficiency refers to a process of progressive deterioration of placental function. It reduces transplacental oxygen and nutrients transfer to the growing fetus, and ultimately limits placental performance during pregnancy [6,7]. However, identifying the molecular mechanisms underlying pathogenesis of placental insufficiency is challenging due to the interference of numerous maternal and genetic confounding factors.

Selective IUGR (sIUGR) is a unique complication of monochorionic (MC) twins in which one twin is an IUGR fetus and the cotwin grows normally. An estimated 10%–15% of MC pregnancies suffer from it [8]. Several studies have extensively investigated the development of sIUGR at the gross histopathological level and have shown that sIUGR is associated with a remarkable discordance of placental sharing wherein the IUGR twin is supported by a smaller portion of the placenta. Also, a higher incidence of abnormal cord insertion occurs in the smaller fetuses [9,10]. Additionally, the terminal branch levels of the placental artery and vein were significantly lower in the IUGR twins [11]. Together, these morphological alterations suggest placental insufficiency and impaired blood perfusion to the fetus in sIUGR. The two fetuses harbor an identical genotype and share the same maternal environment. Thus, sIUGR twins would provide an ideal model to investigate the underlying mechanisms of placental insufficiency by using the healthy cotwin as an internal control to eliminate confounding maternal and genetic factors and to ensure the same gestational age. Recent studies have found that dysregulation of several angiogenesis-related genes, such as *endoglin* and *fms-like tyrosine kinase 1* (*FLT1*), may contribute to the development of sIUGR [12–14]. The *hypoxia inducible factor-1a* (*HIF1A*) mRNA levels, as well as 8-hydroxydeoxyguanosine (8-OHdG) and malondialdehyde (MDA), both biomarkers of oxidative stress, have been demonstrated to be significantly upregulated in IUGR twin placentas, suggesting that oxidative stress might participate in the pathogenesis of sIUGR [15]. In other studies, abnormal placental epigenetic modification, such as altered imprinted genes and DNA methylation, has been found to cause sIUGR [16,17]. However, a systematic placental proteomic profile linked with the functional analysis of clinical sIUGR is still absent.

In this study, we systematically investigated the placental proteome differences between IUGR twins and the normal cotwins utilizing tandem mass tag (TMT)-based technology. In total, we identified 5481 proteins and 233 differentially expressed proteins (DEPs), of which 57 were upregulated and 176 were downregulated. Bioinformatic analysis by ingenuity pathway analysis (IPA) revealed that these DEPs were most significantly enriched in cardiovascular system development and function, as well as organismal survival and organismal development. Notably, 34 DEPs were associated with the biological process of angiogenesis, and we found that angiogenesis was significantly impaired in IUGR twin placentas. The differential expression of *ephrin B2* (*EFNB2*), *vimentin* (*VIM*) and *methionyl aminopeptidase 2* (*METAP2*), which are essential in placental angiogenesis, was validated by PCR and immunohistochemistry analysis. In addition, we found that *metadherin* (*MTDH*) was significantly downregulated in IUGR twin placentas. Knockdown of *MTDH* in human umbilical vein endothelial cells (HUVECs) obviously inhibited the proliferation, migration, tube formation, and spheroid sprouting by affecting numerous genes and biological processes. In brief, our study is the first comprehensive analysis of the placental proteomic profile in sIUGR twins and may offer deeper insights into the molecular mechanisms of IUGR.

## Results

### Clinical characteristics

To identify novel candidate molecules for sIUGR, we harvested 12 placental samples from six sIUGR twin pairs and performed TMT-based comparative proteomic analysis. sIUGR twins, which share the maternal environment and harbor an identical genotype, provide an ideal model for investigating the association between placental development and fetal growth. The demographics and clinical characteristics of the twin pregnancies in this study are presented in Table S1 and Figure S1A–D. All twins were delivered alive by cesarean section at a mean gestational age of 32.3 ± 1.2 weeks. The birth weight of the IUGR twins was, on average, 628 grams lower than that of their healthy cotwins (*P* = 0.0004), with relative birth weight discordance ranging from 25.2% to 48.0%. None of the study population had twin-to-twin transfusion syndrome, fetal malformations, and maternal complications such as preeclampsia or diabetes mellitus. In addition, IUGR twin placentas exhibited more pathological abnormalities, including distal villous hypoplasia, intervillous fibrin deposition, increased syncytial knots, and abnormal cord insertion. (Figure S1E–I).

### Comparative proteomic analysis identifies 233 DEPs in IUGR twin placentas

To comprehensively profile the global placental proteomic changes between IUGR twins and normal cotwins, we applied a high-accuracy quantitative proteomic approach to analyze protein expression (**Figure 1**A). A total of 12 samples from 6 pairs of sIUGR twins were processed in the same experimental period. After protein extraction, reduction, alkylation, and digestion, each 3 pairs of sIUGR twins were labeled with one set of six-plex TMT reagent (group 1: TMT 126, 127, and 128 for normal cotwin 1, 2, and 3; TMT 129, 130, and 131 for IUGR twin 1, 2, and 3 respectively; group 2: TMT 126, 127, and 128 for normal cotwin 4, 5, and 6; TMT 129, 130, and 131 for IUGR twin 4, 5, and 6 respectively). Six isobaric labeled samples within the same set were mixed together and then separated into 10 fractions by high pH reversed-phase liquid chromatography (RPLC). All 20 fractions from two TMT sets were subjected sequentially to liquid chromatography tandem mass spectrometry (LC-MS/MS). MS data were then collected and analyzed for protein identification and quantification. Details on MS data analysis were presented in the Methods section. Specifically, we identified a total of 5481 nonredundant proteins in sIUGR twin placentas (Table S2). A histogram plot revealed the fold change (FC) distribution of all identified proteins (Figure 1B). The distribution of statistical significance (−log_10_ transformed *P* value) and degree of change (log_2_ transformed FC) for all identified proteins were presented in the volcano plot (Figure 1C). Using a combination of *P* < 0.05 and |FC| > 1.3, 233 proteins displayed significant differential expression in IUGR twin placentas compared to the normal cotwin placentas. Of these, 57 DEPs were upregulated and 176 DEPs were downregulated (Table S3). Unsupervised hierarchical clustering of the DEPs revealed good separation between the two groups (Figure 1D).

**Figure 1.**
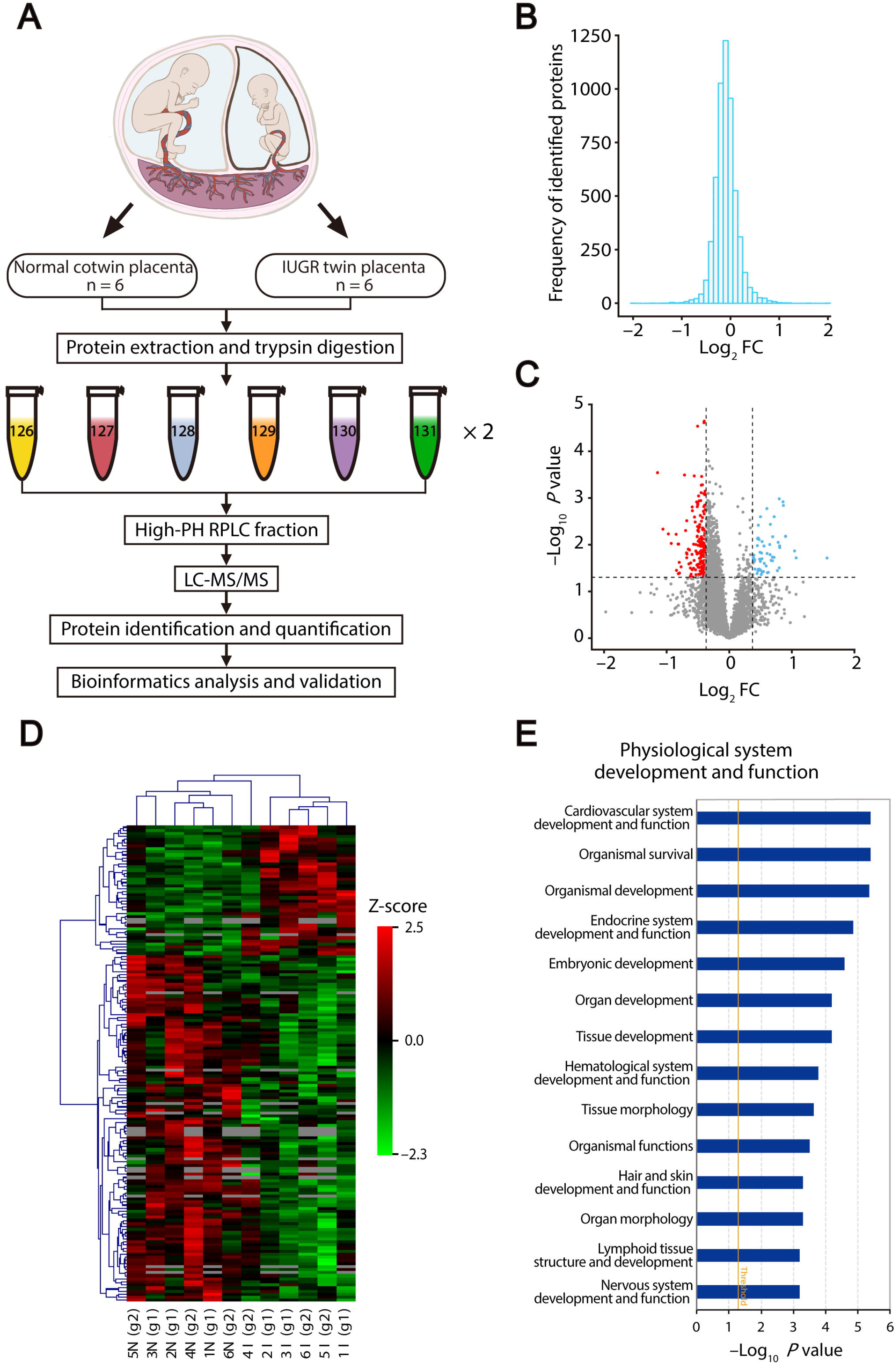
Identification and biofunction analysis of DEPs in IUGR twin placentas. **A**. Proteomic workflow for placental tissues from normal cotwins and IUGR twins. Twelve samples from 6 sIUGR pregnancies were enrolled in this study and labeled with two sets of six-plex TMT reagents. **B**. Histogram plot showing the log_2_ FC distribution of all identified proteins. **C**. Volcano plot of DEPs between IUGR twin placentas and normal cotwin placentas. DEPs were selected with *P* < 0.05 and |FC| > 1.3. Red spots indicate the downregulated DEPs, blue spots represent the upregulated DEPs, and gray spots indicate proteins without significantly different expression. **D**. Heatmap of the DEPs identified in IUGR twin placentas. Proteins are represented by rows, and samples by columns. **E**. Functional analysis of the DEPs in IUGR twin placentas using IPA software. DEPs, differentially expressed proteins; IUGR, intrauterine growth restriction; sIUGR, selective IUGR; RPLC, reverse-phase liquid chromatography; FC, fold change; LC-MS/MS, liquid chromatography tandem mass spectrometry; N, normal cotwin; I, IUGR twin; TMT, tandem mass tag; IPA, ingenuity pathway analysis.

To deepen the biological insights into these DEPs, we utilized IPA software for bioinformatics analysis. The core biofunction analysis of these DEPs was divided into three categories. In “physiological system development and function” category, the most significantly enriched item was “cardiovascular system development and function”, which involved 52 DEPs. The other two main enriched subcategories were “organismal survival” (number of DEPs in this function [n] = 66) and “organismal development” (n = 71) (Figure 1E; Table S4). In addition, in “molecular and cellular function” category, the top three enriched functions included “cell death and survival”, “lipid metabolism”, and “molecular transport” (Figure S2A; Table S4). For “diseases and disorders”, the DEPs were significantly related to “endocrine system disorders”, “gastrointestinal disease”, and “metabolic disease” (Figure S2B; Table S4). Additionally, IPA canonical pathway analysis identified oxidative phosphorylation, mitochondrial dysfunction, and the Sirtuin signaling pathway as the top three pathways that might be potentially affected in IUGR twin placentas (Figure S2C).

### Angiogenesis is significantly impaired in IUGR twin placentas

Among the 52 DEPs involved in “cardiovascular system development and function”, 34 DEPs were enriched in the biological process of angiogenesis encompassed in this biofunction, and the state of angiogenesis was predicted to be suppressed by IPA (**Figure 2**A). Placental angiogenesis is crucial to villous development and placental function. Any impairment in placental angiogenesis can lead to poor placentation, decreased blood flow to the fetus, and various pregnancy complications [18]. To confirm whether angiogenesis was altered in IUGR twin placentas, we assessed the microvessel density (MVD) and vascular area density per sample following immunostaining of CD34 as an endothelial cell marker. A significant decrease in MVD was present in IUGR twin placentas, while the vascular area density was also significantly lower in IUGR twin placentas than in the normal cotwins (Figure 2B–D). Taken together, these data suggested that impaired placental angiogenesis in IUGR twins might be responsible for the development of sIUGR. To further validate the proteomic results, we selected 3 markers (*EFNB2, VIM*, and *METAP2*) for immunohistochemistry analysis. The 3 markers were reported to play essential roles in placental angiogenesis and fetal development [19–21]. As shown in Figure 2E–H, the intensity of VIM, EFNB2, and METAP2 immunoreactivity was significantly lower in IUGR twin placentas than in normal cotwins, which was in line with TMT proteomic results. Additionally, we performed qRT-PCR to explore their expression changes at mRNA level. Similar to the proteomic data, *VIM, EFNB2*, and *METAP2* mRNA expression levels were significantly downregulated in IUGR twin placentas (Figure 2I–K).

**Figure 2.**
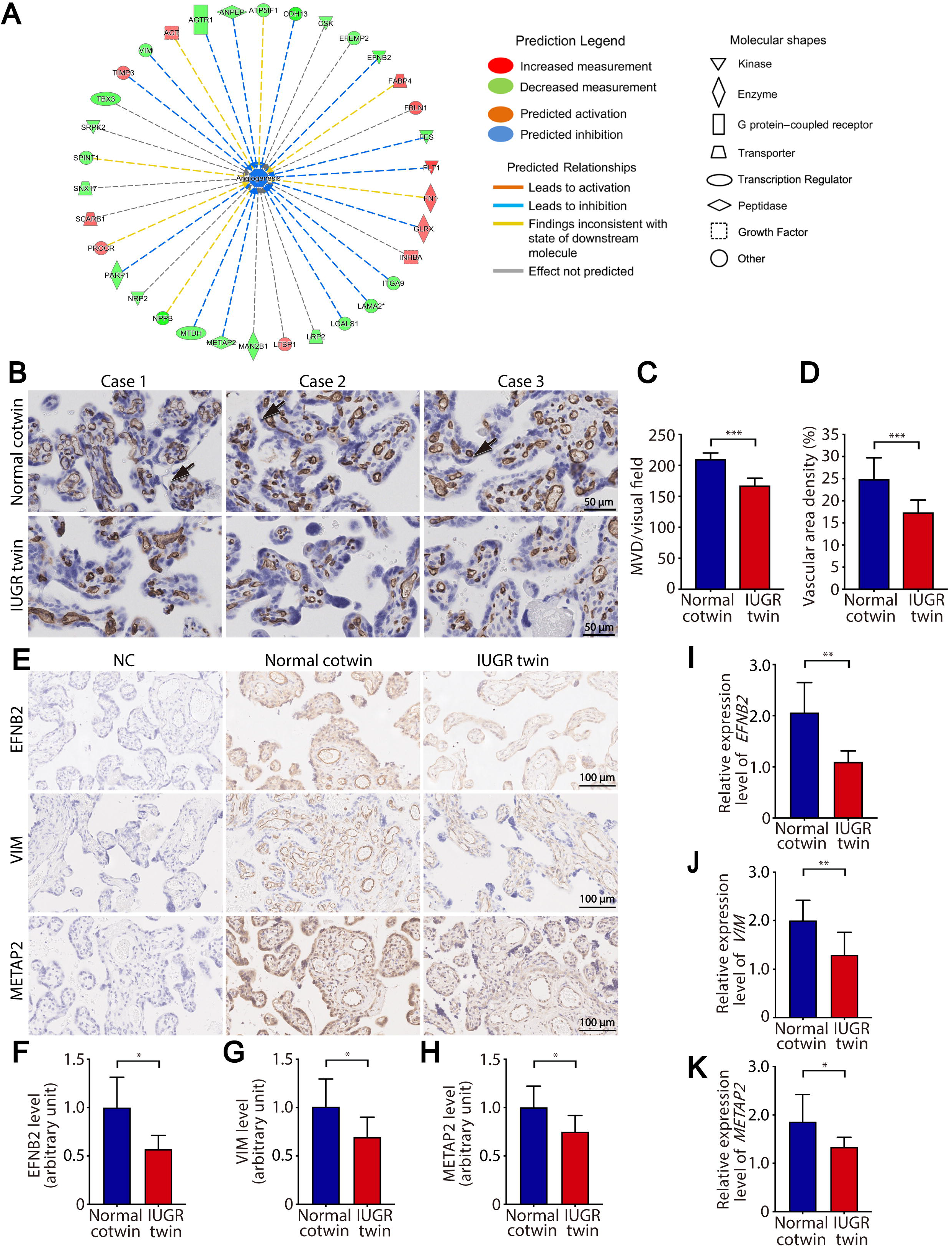
Impairment of angiogenesis in IUGR twin placentas. **A**. IPA identified the “angiogenesis” annotation to be affected in IUGR twin placentas.**B**. Immunostaining of CD34 in human placentas from IUGR twins and normal cotwins. CD34 is limited to endothelial cells. Scale bar, 50 μm. **C**. Placental MVD in IUGR twins and normal cotwins. **D**. Placental vascular area density in IUGR twins and normal cotwins. **E**. Immunohistochemical images of EFNB2, VIM, and METAP2 in placental shares from IUGR twins and normal cotwins. Scale bar, 100 μm. **F**–**H**. EFNB2, VIM, and METAP2 immunostaining intensity per tissue area is represented by the fold-decrease relative to the normal control mean. **I**–**K**. qRT-PCR analysis showed that the *EFNB2, VIM*, and *METAP2* transcriptional levels were significantly lower in placentas of IUGR twins compared with normal cotwins. All data are shown as mean ± SD with 6 samples per group. *, *P* < 0.05; **, *P* < 0.01; ***, *P* < 0.001. MVD, microvessel density; NC, negative control; SD, standard deviation.

### *MTDH* silencing inhibits HUVEC proliferation and migration

Among the 34 DEPs related to angiogenesis, we also observed a significant decrease in *MTDH* expression in IUGR twin placentas (**Figure 3**A–C). There is evidence that *MTDH* participates in tumor angiogenesis [22,23], while its role in placental angiogenesis remains unclear. During placental angiogenesis, endothelial cells undergo proliferative, migratory, and morphological events to form a vessel network. To investigate the causality between endothelial *MTDH* deficiency and sIUGR development, the role of *MTDH* in the proliferation and migration of HUVECs was explored. HUVECs were transfected with negative control siRNA (si-NC) and siRNA against *MTDH* (si-*MTDH*). Transfection with si-*MTDH* markedly decreased *MTDH* mRNA and protein levels (Figure 3D–F). The proliferation and migration abilities of HUVECs were examined using an electrical impedance-based tool, the xCELLigence® real-time cell analysis (RTCA) system, which enables continuous monitoring of cell behaviors and allows for real-time assessment of multiple cell functions. For the proliferation assay, cells treated with si-NC or si-*MTDH* were seeded in the wells of an E-plate. Cells were cultured for 60 h, and the si-*MTDH* group exhibited a significant reduction in proliferation ability (Figure 3G). Cell migration was assessed using a CIM-16 plate with upper and lower chambers in each well. The results revealed that *MTDH* interference obviously decreased HUVEC migration ability compared with that of control cells (Figure 3H). To further confirm the effect of *MTDH* deficiency on HUVEC migration in the horizontal direction, we performed a wound healing assay. As expected, the wound in the si-*MTDH*-transfected cell culture healed more slowly than that of the control group (Figure 3I, J). In summary, these findings demonstrated that *MTDH* is vital to HUVECs proliferation and migration.

**Figure 3.**
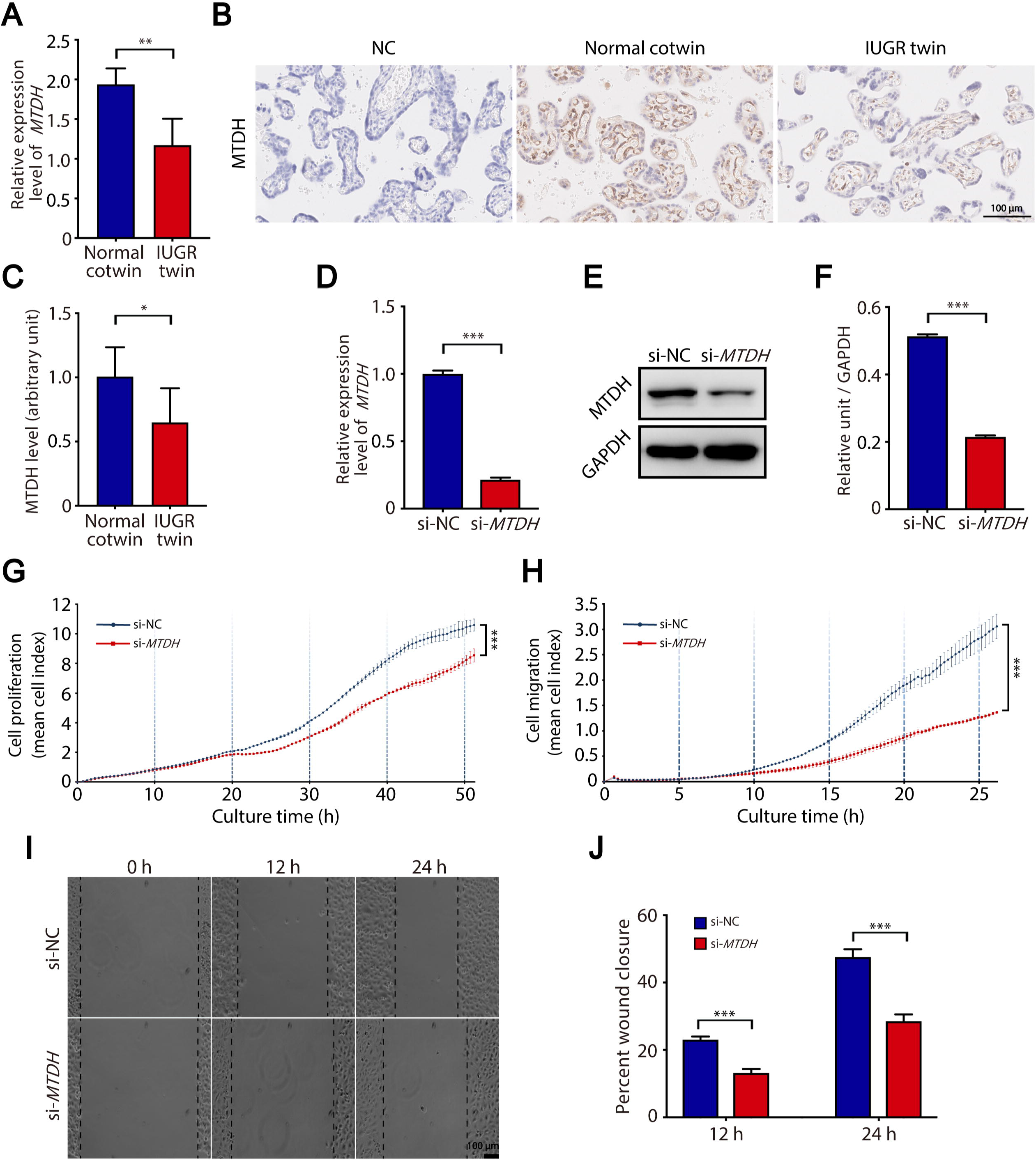
*MTDH* expression is downregulated in IUGR twin placentas, and *MTDH* knockdown impairs HUVECs proliferation and migration. **A**. *MTDH* mRNA levels in IUGR twin placentas (n = 6) and normal cotwin placentas (n = 6) by qRT-PCR analysis. **B**. MTDH expression in placental shares of IUGR twins and normal cotwins by immunohistochemistry. Scale bar, 100 μm. **C**. MTDH immunostaining intensity in IUGR twin and normal cotwin placentas (n = 6 in each group). **D**–**F**. After transfection for 48 h, the interference efficiency of siRNA targeting *MTDH* in HUVECs was evaluated by qRT-PCR (**D**) and western blotting (**E**). The relative intensity of MTDH levels was evaluated by ImageJ from three independent experiments (**F**). **G**. Proliferation curves showed that *MTDH* knockdown greatly inhibited HUVECs proliferation. Two-way ANOVA was used for data analysis. **H**. Migration curves revealed that *MTDH* knockdown greatly inhibited HUVECs migration. Two-way ANOVA was used for data analysis. **I**–**J**. Knockdown of *MTDH* in HUVECs decreased the rate of wound closure. Scale bar, 100 μm. All experiments had 3 replicates with results shown as mean ± SD. *, *P* < 0.05; **, *P* < 0.01; ***, *P* < 0.001. ANOVA, analysis of variance.

### Knockdown of *MTDH* inhibits tube formation and spheroid sprouting in HUVECs

To further determine *MTDH*’s effect on placental angiogenesis, the 2-dimensional (2D) tube formation assay and 3-dimensional (3D) spheroid sprouting assay were conducted using HUVECs. As shown in **Figure 4**A, si-NC-transfected HUVECs could form capillary-like structures in a few hours on Matrigel, while the tube formation capacity was perturbed in the si-*MTDH* group. *MTDH* deficiency distinctly impaired the number and area of tubes, the total tube length, as well as the number of branching points (Figure 4B–E). Similarly, when embedded in 3D collagen I gels, spheroids composed of si-NC-transfected HUVECs retained the ability to produce capillary-like outgrowths, while only a few sprouts were formed in *MTDH*-depleted HUVEC spheroids (Figure 4F). Compared to the control cells, the si-*MTDH* HUVECs exhibited a significant reduction in cumulative sprout length (Figure 4G). Accordingly, these data suggest that downregulation of *MTDH* inhibits placental angiogenesis.

**Figure 4.**
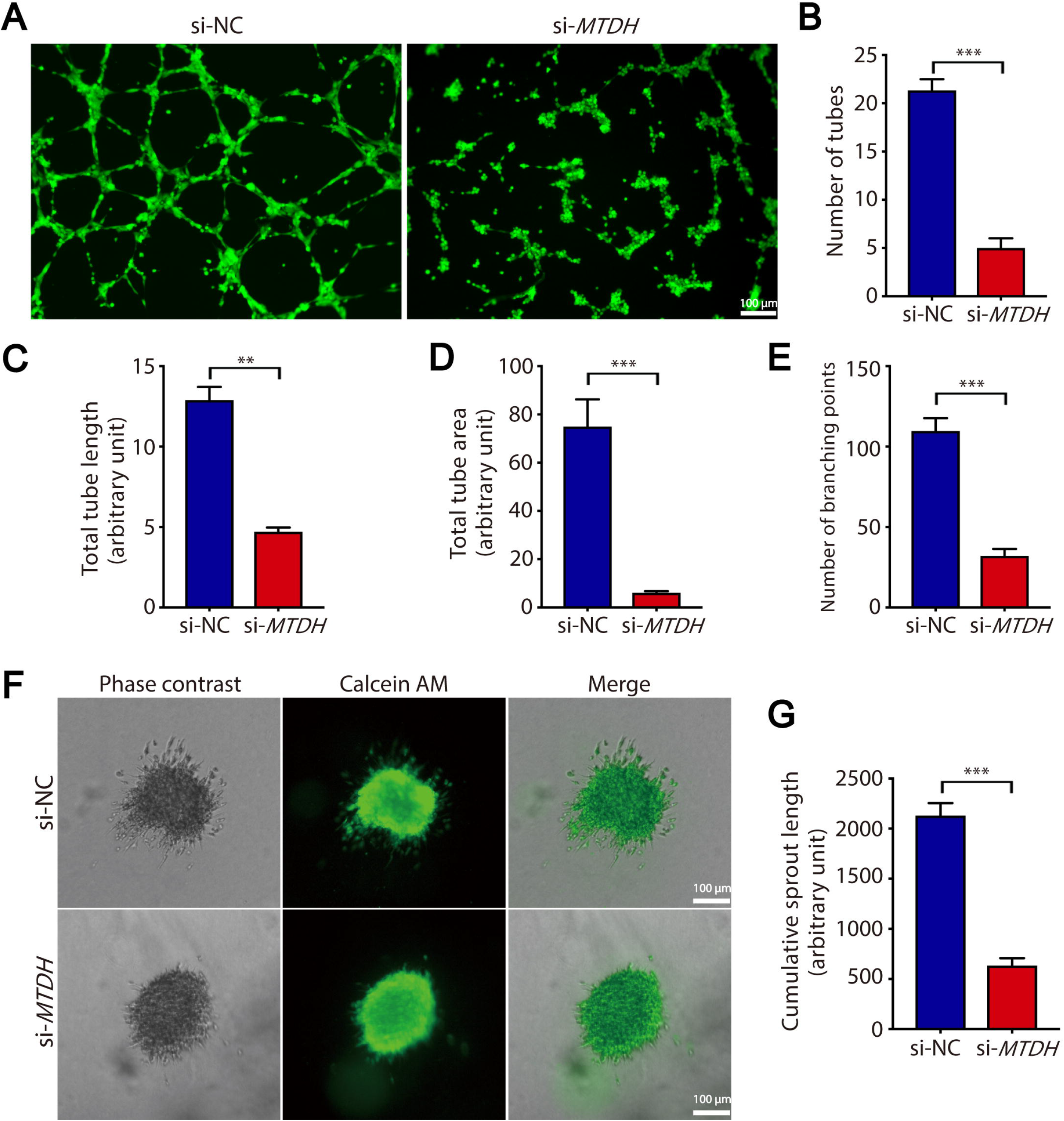
*MTDH* knockdown inhibits HUVECs tube formation and spheroid sprouting. **A**. HUVECs were harvested 48 h after transfection with si-NC or si-*MTDH* and then plated in Matrigel-coated wells. After 8 h, HUVEC tube formation was observed under a LeicaDMI4000B microscope and photographed. Scale bar, 100 μm. **B**–**E**. Quantification of the number of tubes (**B**), total tube length (**C**), total tube area (**D**), and number of branching points (**E**) in the si-*MTDH* group and the si-NC group using ImageJ software. The results were from three independent assays. **F**. si-NC or si-*MTDH* transfected HUVECs spheroids were photographed 48 h later after embedding in collagen gels. Scale bar, 100 μm. **G**. ImageJ software was used to quantify the cumulative sprout length in the si-*MTDH* group and the si-NC group. All data are expressed as mean ± SD from 3 experiments. **, *P* < 0.01; ***, *P* < 0.001.

### *MTDH* is required in placental development and fetal growth

To verify whether *MTDH* is required in placental and fetal development, we knocked down *MTDH* in HUVECs and performed RNA-seq analysis. Both the control and si-*MTDH* groups had three biological replicates each. The correlation analysis based on the overall RNA-seq data for each sample pair showed that the correlation coefficient between intra-group samples was higher than that between inter-group samples, indicating that samples within groups had strong resemblance in terms of transcription pattern and the biological experimental operations were reliable and reproducible, while the gene expression changed following MTDH interference. (**Figure 5**A). The top 50 protein-coding genes with significantly differential expression (adjusted *P* < 0.05) were selected as candidates for further analysis (Table S5). Among the 50 genes, 29 were upregulated and 21 were downregulated in the si-*MTDH* group. The expression profiles of the 50 genes are presented in a heatmap (Figure 5B). Further functional enrichment analysis revealed that 50 candidate genes were associated with various biological processes, such as defense responses, apoptotic processes, actin filament organization, negative regulation of cell proliferation, and angiogenesis (Figure 5C). The interaction network of the 50 candidates is presented in Figure 5D.

**Figure 5.**
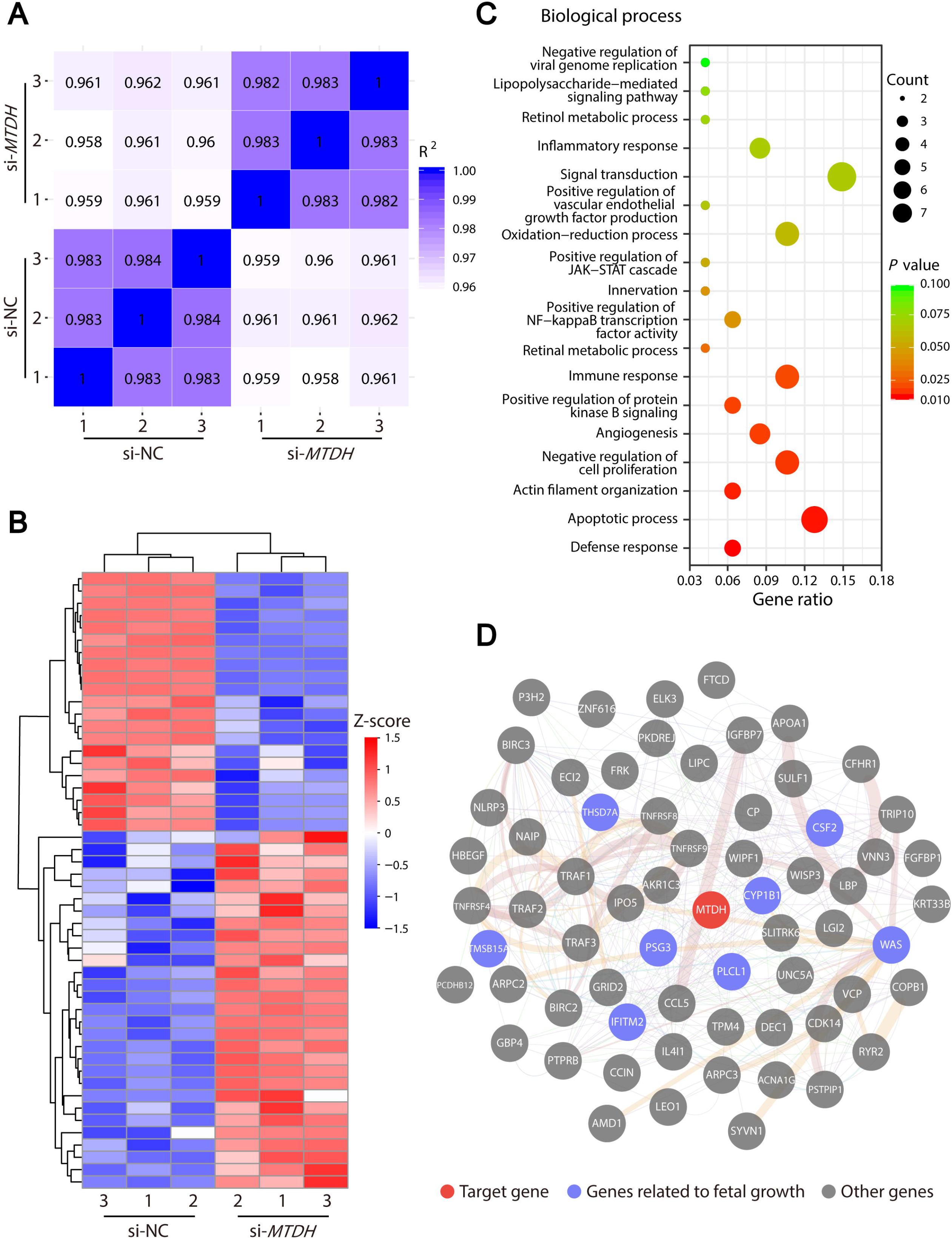
RNA-seq analysis of HUVECs following *MTDH* interference. **A**. Correlation coefficient analysis based on RNA-seq data for each sample pair. Three replicates per condition. **B**. Heatmap of the top 50 differentially expressed protein-coding genes between si-*MTDH* group and si-NC group. Genes are represented by rows, and samples by columns. **C**. Bubble plot depicting biological process enrichment for 50 candidates by gene ontology. **D**. Interaction network of the 50 candidate genes. The network was constructed by GeneMANIA and visualized using Cytoscape 3.7.2.

Among the 50 candidates, we found quite a few genes previously reported to be implicated in placental dysfunction and fetal growth restriction, including *PSG3* (upregulation of *PSG3* in both maternal blood and IUGR placenta) [24,25], *CSF2* (*Csf2*-deficient *mice* exhibit placental insufficiency and fetal growth retardation) [26,27], *THSD7A* (higher expression of *THSD7A* is associated with low birth weight) [28], *CYP1B1* (lower methylation levels of *CYP1B1* are found in preterm delivery with reduced fetal growth) [29], and *PLCL1* (double-knockout *mice* for *Plcl1* and *Plcl2* exhibit reduced litter events and litter size) [30]. These results further provide support for the validity of our results. Interestingly, we also found several unreported genes that may affect placental development and fetal growth, such as *IFITM2, TMSB15A*, and *WAS*. These findings may help further the discovery of potential biomarkers and therapeutic targets.

## Discussion

Despite advances in perinatal care, IUGR remains a challenging problem faced by obstetricians worldwide. Placental insufficiency and subsequent undernutrition and hypoxia are considered to be the ultimate pathomechanisms that disturb fetal growth [31]. However, the molecular regulatory mechanisms of IUGR have not been fully elucidated due to genetic and maternal heterogeneity. In monochorionic twin pregnancy, the IUGR twin and the normal cotwin have an identical genotype and develop in the same uterus, making it an excellent model to investigate placental disorders [32]. Proteins, which act as the ultimate executors of gene function, are vital players within organisms in terms of functionality and structure [33]. Thus, investigating the molecular basis for IUGR at the protein level is of paramount importance to shed light on its clinical diagnosis and treatment. Based on the above information, we sought to construct a comprehensive profile of the placental DEPs by taking advantage of sIUGR twins. In this work, we performed TMT-based comparative proteomic studies using placental samples of six sIUGR twin pairs from the clinic. A total of 5481 unique proteins with high confidence were detected in human placental tissues. Therein, 233 proteins were identified as DEPs between IUGR twin and normal cotwin placentas, with 57 upregulated and 176 downregulated. In the current work, numerous proteins relevant to embryonic development were found to be dysregulated in IUGR twin placentas. As the interface of interaction and communication between mother and fetus, the placenta is essential to embryo development and, in particular, to fetal growth [34]. The aberrant expression of these proteins in IUGR twin placentas implies that they may play key roles in impairing fetal growth potential. In addition, a set of proteins that can modulate lipid metabolism were detected in our study. Over the past few decades, lipid metabolism during pregnancy has been highlighted as essential not only for optimal placental development and function but also for proper intrauterine growth of the fetus [35]. Disrupted lipid metabolism may contribute to placental insufficiency and suboptimal fetal growth, which is consistent with previous discoveries in singleton IUGR cases [36,37]. In addition, proteins that are key players in molecular transport appeared prominently among the top functional terms. In fact, the formation of an efficient transport interface is the essence of the placenta, which allows the growing fetus to fulfill its developmental demands from maternal circulation [34]. Alterations in nutrient transporter expression have also been evidenced in IUGR placentas [38,39].

Notably, we found that 34 DEPs were implicated in angiogenesis. Adequate angiogenesis ensures that placentas develop as highly vascularized organs, thereby supporting the blood flow required for fetal growth [40]. Aberrant placental vasculature has been identified in nearly all human pregnancy complications, further confirming the importance of placental angiogenesis for normal pregnancy [41]. A previous study discovered significantly decreased villus vascular density in placentas of singleton IUGR [42]. Our results also confirmed that placental MVD and vascular area density were significantly lower in smaller fetuses. These results together suggest that impairment of placental angiogenesis may play key roles in the incidence and development of IUGR. *MTDH*, also known as *AEG-1*, is implicated as an oncogene [43,44]. According to our findings, *MTDH* was significantly downregulated in IUGR twin placentas and located in intravillous capillary endothelial cells. In addition, we verified that the function of *MTDH* is critical in placental angiogenesis, indicating that decreased expression of *MTDH* may contribute to the pathogenesis of placental dysfunction and fetal growth retardation. While our understanding of the molecular mechanisms of IUGR has advanced significantly over the past few decades, there are currently no reliable diagnostic biomarkers or targeted therapies available for this disease in clinical practice [45,46]. Translational application of the candidate targets identified in singleton IUGR pregnancies is very limited, probably mainly due to the heterogeneity of the samples in these studies. Thus, MTDH identified in our investigation using sIUGR twins may serve as a potential biomarker for early prediction and accurate diagnosis of IUGR, as well as a novel target for future treatment. In summary, our findings comprehensively described the proteomic landscape of sIUGR twin placentas, which may provide reliable resources for mechanistic studies of the core regulators involved in placental dysfunction and fetal growth restriction.

## Materials and methods

### Patients and clinical specimen collection

Placental samples from 6 sIUGR twins were collected at Peking University Third Hospital. Ultrasonography at 11–14 weeks was conducted to determine the chorionicity, and postnatal placental examination was used to confirm it. sIUGR refers to a monochorionic twin in which the estimated fetal weight (EFW) of one fetus is lower than 10th percentile while the cotwin’s is normal and the intertwin EFW discordance is more than 25%. EFW was calculated based on Hadlock formula [47]. EFW discordance was calculated with this formula: (normal cotwin EFW−IUGR twin EFW)/normal cotwin EFW. All participants underwent cesarean delivery, and the samples were collected within 30 min. Samples were obtained from the maternal side of the placental lobules within 2 cm of each umbilical cord insertion site, aiming to reduce possible bias caused by the regional variations in placental gene expression associated with the sampling sites [48,49]. The placental biopsies were excised in areas free of macroscopic lesions of calcification or infarction after removing the maternal deciduas and fetal membranes. The biopsy was washed in PBS for blood removal. A portion of the sample was stored at −80°C after freezing in liquid nitrogen. The rest was fixed in 10% formalin solution followed by embedding in paraffin.

### Protein extraction, digestion, and TMT labeling

Placental tissues were lysed using 7 M urea with 1% (w/v) dithiothreitol (DTT), 2 M thiourea, and 1% (v/v) protease inhibitor cocktail. Equal amounts of protein (100 µg/sample) were used for subsequent processing. 1 M DTT was used for peptide reduction at 56°C for 30 min and 1 M iodoacetamide for alkylation at room temperature for 30 min in darkness. Samples were diluted with 25 mM (pH 8.2) Tris-HCl and 0.1 M CaCl2 until the final concentration of urea was 1 mM. Trypsin (Catalog No. V5111, Promega, Madison, WI) was used to digest proteins for 12 h. The resulting peptides were desalted on Sep-Pak C18 columns (Waters, Altrincham Road, Wilmslow, UK), and then labeled with two sets of six-plex TMT reagents (Catalog No. 90062, Thermo Fisher Scientific, Waltham, MA). Each 6 isobaric labeled samples within the same set were combined and vacuum dried for further analysis.

### LC-MS/MS

The peptide mixture was separated into 10 fractions by high pH RPLC using BEH C18 columns (1.7 µm, 1.0 mm × 150 mm) attached to the H-Class System (Waters). The second-dimensional liquid chromatography separation of the peptides was conducted on the Ekspert™ nano LC 415 instrument (AB Sciex, Toronto, Canada). The peptides were eluted using 2% acetonitrile with 0.1% formic acid as buffer A and 98% acetonitrile with 0.1% formic acid as buffer B with a gradient of 100 min. The eluted peptides were subsequently analyzed using TripleTOF 5600+ mass spectrometer (AB Sciex). Data were obtained in positive ion mode. For TOF-MS scan, the mass range was set as 350–1500□*m/z*. The MS/MS scan was carried out with a mass range of 100–1500□*m/z*.

### Data analysis

The raw data were analyzed by using the MaxQuant software (Version 1.5.2.8) [50] and the human UniProt reference protein sequences [51] to generate initial spectra-peptide matches and quantification values. The search was performed with the following parameters: enzyme, trypsin/p; max missed cleavages, 2; 0.1 Da for first search mass tolerance; 0.01 Da for main search mass tolerance; variable modifications, acetylation (N-term) and oxidation (M); fixed modification, carbamidomethyl (C). Protein identification requires at least one unique peptide. False discovery rate (FDR) was 0.05 for both peptide and protein identification. Then, the MaxQuant results were further processed by the MaxReport software [52] to correct and optimize the quantitative results. In brief, the reporting intensity of each isobaric label was corrected by an impurity correction matrix. Additionally, each label (sample) was normalized by the total intensity. Then, the relative protein expressions were calculated by the Libra algorithm [53]. Finally, DEPs were determined by *P* < 0.05 and |FC| > 1.3. The MaxReport quantification workflow was also successfully tested by using a benchmark dataset (PXD000001) to quantify the spiked reference proteins. Additionally, all of the samples were harvested and processed in the same experimental period. Due to the limited number of TMT labels, these samples were labeled separately and loaded into the mass spectrometer in a sequential process.

Comparison of the average intensities and unique peptides between the two six-plex TMT experiments indicated that there was a minor batch effect between the two groups, as the Pearson correlation coefficient values for intensities and peptides were 0.96 and 0.98, respectively. Considering only two sequential batches in this study, we did not perform such correction. The global protein expression patterns of sIUGR twin placentas were illustrated using histograms and volcano plots in the R package (version 3.5.2). Hierarchical cluster analysis of the DEPs was performed with MeV software (version 4.9.0). We used IPA software (QIAGEN, Redwood City, CA) to identify the biofunctions that were significantly associated with the DEPs. Specifically, the list of DEPs, containing gene symbols, fold changes and *P* values, was uploaded into IPA software for core analysis. The core analysis included enrichment of diseases and functions, canonical pathways, and networks. The core analysis was conducted with the setting of direct and indirect relationships between focused genes based on the human section of “Ingenuity Knowledge Base”, which was a manually curated database of biological interactions and functional annotations.

### Immunohistochemistry

After deparaffinization, antigen retrieval and endogenous peroxide blocking, the sections (5 µm in thickness) were incubated with primary anti-CD34 antibody (Catalog No. ZM-0046, ZSGB-BIO, Beijing, China), anti-EFNB2 antibody (1:50; Catalog No. HPA008999, Atlas Antibodies, Bromma, Sweden), anti-METAP2 antibody (1:3000; Catalog No. HPA019095, Atlas Antibodies), anti-VIM antibody (Catalog No. ZM-0260, ZSGB-BIO) and anti-MTDH antibody (1:150; Catalog No. ab124789, Abcam, Cambridge, UK). After incubating with secondary antibodies, the sections were stained with diaminobenzidine solution (Catalog No. ZLI-9018, ZSGB-BIO). The expression level of the target genes was assessed with Image-Pro Plus (Media Cybernetics, Rockville, MD).

### MVD and vascular area density

Using CD34 as an endothelial cell marker, the MVD was used to evaluate placental angiogenesis by measuring the immunostaining of CD34 in placental sections. Briefly, the number of CD34-positive vessels was counted under a magnification of 400× in 5 nonconsecutive fields by two observers who were blinded to the control and case groups. The mean value of the 5 fields was expressed as the MVD. The vascular area density was evaluated as follows: 5 images per slide with a magnification of ×200 were captured by NanoZoomer-SQ (Catalog No. C13140-01, Hamamatsu, Shizuoka, Japan) and NDP.view2 software (Hamamatsu). Image-Pro Plus (Media Cybernetics) was used for measurement of the vascular area and the villous area. The mean of the percentage of vascular area in villous area in 5 images was defined as the vascular area density.

### RNA extraction and qRT-PCR

RNA was extracted using TRIzol (Catalog No. 15596018, Thermo Fisher Scientific), and then reverse-transcribed into cDNA. qRT-PCR was conducted with the SYBR™ Green Master Mix reagent (Catalog No. A25742, Thermo Fisher Scientific). Internal reference gene was *glyceraldehyde-3-phosphate dehydrogenase* (*GAPDH*). All experiments were replicated three times independently. Sangon Biotech Corporation (Shanghai, China) supplied the primers (Table S6).

### Cell culture

Immortalized HUVECs were a generous gift from Peking University Third Hospital Medical Research Center. HUVECs were cultured in DMEM/F-12 medium (Catalog No. C11330500BT, Thermo Fisher Scientific) containing 1% penicillin/streptomycin (Catalog No. V900929, Sigma-Aldrich, St. Louis, MO) as well as 10% fetal bovine serum (FBS) (Catalog No. 10099141, Thermo Fisher Scientific) at 37°C with 5% CO_2_. HUVECs were authenticated with short tandem repeat analysis.

### Cell transfection

Transient transfection of HUVECs with siRNA was performed at 70% confluence in six-well plates using Lipofectamine 3000 (Catalog No. L3000015, Thermo Fisher Scientific). *MTDH* siRNA sequences were as follows: sense, (5’–3’) GGAACCAAUUCCUGAUGAUTT; antisense (5’–3’) AUCAUCAGGAAUUGGUUCCTT. The siRNA was synthesized by GenePharma (Shanghai, China).

### Western blotting

Proteins (20 mg for each sample) were electrophoresed on 8% SDS-PAGE gels and then transferred to polyvinylidene fluoride membranes (Catalog No. IPVH00010, Millipore Bedford, MA). The membranes were blocked in 5% nonfat dry milk followed by incubating with anti-MTDH antibody (1:10,000; Catalog No. ab124789, Abcam) and anti-GAPDH antibody (1:5000; Catalog No. ab8245, Abcam) overnight at 4°C. After incubating with secondary antibodies, the bands were revealed with an enhanced ECL Kit (Catalog No. BF06053-500, Biodragon, Beijing, China) and analyzed by ImageJ (National Institutes of Health, NIH) using GAPDH as the loading control.

### Real-time cell proliferation and migration assays

The RTCA analyzer (ACEA Biosciences, San Diego, CA) was used to examine cell proliferation and migration abilities. The cell proliferation assay was carried out on the E-plate with each well consisting of 5□×□10^3^ cells treated with si-*MTDH* or si-NC. The impedance of each well, expressed as the cell index (CI), was automatically measured every 30 min for 60 h. The migration assay was performed in a CIM-16 plate. A total of 2□×□10^4^ transfected cells were seeded into the upper chamber with serum-free medium, while it was complete medium in the lower chamber. Scans were run with sweeps every 15 min for 60 h. The impendence of each well was also represented by the CI.

### Wound healing assay

In 6-well plates, 5 × 10^5^ HUVECs transfected with either si-*MTDH* or si-NC were seeded per well. When cells reached confluence, scratches were made with 200-µl pipette tips. Afterwards, serum-free medium was used for cell culture, and images were captured every 12 h for a total of 24 h. The NIH ImageJ software was applied to analyze the cell migration distance.

### Tube formation assay

In 96-well plates, 60 µl of Matrigel (Catalog No. 356234, BD Biosciences, Bedford, MA) was placed in each well, which was then solidified at 37°C for 30 min. Transfected HUVECs (2 × 10^4^/well) were seeded into Matrigel-coated wells and cultured for 8 h. After that, the medium was discarded, and the cells were labeled with 2 µM calcein AM fluorescent dye (Catalog No. ab141420, Abcam) at 37°C for 20 min. The LeicaDMI4000B inverted microscope was used to observe the capillary-like structures, and images were taken using a Leica DFC345FX camera. The number and area of the formed tubes, the number of junctions, as well as the total branching length were quantified using ImageJ software (NIH). Experiments were performed 3 times independently.

### Spheroid sprouting assay

As previously described, we performed the 3D sprouting assay [54]. HUVECs were harvested after 24 h of transfection with either si-*MTDH* or si-NC to generate endothelial cell spheroids. Briefly, HUVECs (1 × 10^3^/well) were added into nonadherent round-bottom 96-well plates, and the cells spontaneously formed a single EC spheroid in each well within 24 h. The spheroids were then embedded in collagen gel and cultured for 48 h. A LeicaDMI4000B inverted microscope was used to observe the spheroids after labeling with calcein AM. ImageJ software (NIH) was used to measure all sprouts originating from each spheroid. The cumulative sprout length of 10 spheroids were compared between the 2 groups. The experiments were replicated 3 times independently.

### RNA-seq and data analysis

RNA was isolated and quality controlled. The cDNA libraries of transfected HUVECs were constructed and purified, followed by sequencing on Illumina NovaSeq platform. Clean reads were achieved by processing raw data with in-house Perl scripts. HISAT2 was used for reads alignment and featureCounts for counting reads. DESeq2 was applied for differentially expressed genes identification (adjusted *P* < 0.05). DAVID v6.8 was used for gene ontology (GO) analysis. GeneMANIA was carried out to construct protein-protein interaction network.

### Statistical analysis

Proliferation and migration curves were estimated by two-way ANOVA using SPSS software version 25.0. Paired *t*-test was applied to compare the differences between IUGR twins and the normal cotwins. All other data comparisons were conducted with Student’s *t*-test. *P* < 0.05 was considered statistically significant.

## Ethical statement

Ethical approval of our study was achieved from the Ethics Committee of the Peking University Third Hospital (IRB00006761-2016145). Prior to placental samples collection, written informed consent was obtained from participants.

## Data Availability

All data produced are available online at iProX database and Genome Sequence Archive.

https://www.iprox.cn/page/project.html?id=IPX0004712000

https://ngdc.cncb.ac.cn/gsa-human/browse/HRA002656

## Data availability

The placental proteomic data of sIUGR twins have been uploaded to the iProX database (https://www.iprox.org/) [55], and are accessible with the dataset identifier IPX0004712000. The RNA-seq data of HUVECs have been submitted to the Genome Sequence Archive (GSA: HRA002656) at https://ngdc.cncb.ac.cn/gsa-human/ [56].

## Acknowledgments

Our investigation was supported by the National Natural Science Foundation of China (Grant Nos. 81971399 and 82171661).

## Supplementary material

**Figure S1 Clinical characteristics and placental histopathology of sIUGR twins**

**A**–**B**. Representative images of ultrasound biometric measurements of the normal cotwin (**A**) and the IUGR twin (**B**) from a sIUGR pregnancy, including BPD, HC, AC, and FL. **C**. Ultrasound estimated fetal weight of the normal cotwins and the IUGR twins from six pairs of sIUGR pregnancies throughout gestation were plotted on the fetal growth curve. **D**. Birth weight of the normal cotwins and the IUGR twins (n = 6 in each group). **E**. Normal placental parenchyma from the normal cotwin. Scale bar, 100 μm. **F**. Distal villous hypoplasia in the IUGR twin placenta. Scale bar, 100 μm. **G**. Increased fibrin deposition (arrow) in the IUGR twin placenta. Scale bar, 100 μm. **H**. Increased syncytial knots (arrow) in the IUGR twin placenta. Scale bar, 100 μm. **I**. A representative image of sIUGR twin placenta after dye injection. Placental sharing was indicated by the white dotted line. Velamentous cord insertion was observed in the IUGR twin. ***, *P* < 0.001. BPD, biparietal diameter; HC, head circumference; AC, abdominal circumference; FL, femur length.

**Figure S2 Bioinformatic analysis of the DEPs in IUGR twin placentas by IPA software**

**A**. DEPs significantly enriched in the “molecular and cellular function” category. **B**. DEPs significantly enriched in the “disease and disorder” category. **C**. Canonical pathway analysis of the DEPs.

**Table S1 Clinical features of 6 sIUGR twins**

**Table S2 Detailed information on 5481 nonredundant proteins identified in sIUGR twin placentas**

**Table S3 Detailed information on 233 DEPs in IUGR twin placentas**

**Table S4 Detailed information on the biofunction analysis by IPA**

**Table S5 The top 50 differentially expressed protein-coding genes between the si-*MTDH* group and the si-NC group**

**Table S6 Primers used for qRT-PCR**

