## Supplementary material for "The Proteome Landscape of Human Placentas for Monochorionic Twins with Selective Intrauterine Growth Restriction": Table S1.docx

| **Table S1 Clinical features of 6 sIUGR twins** | |
| --- | --- |
| **Characteristic** | **Value** |
| Maternal age (years) | 32.8 ± 3.8 |
| Nulliparous | 2 (33.3%) |
| Mode of conception |  |
| Spontaneous | 5 (83.3%) |
| IVF-ET | 1 (16.7%) |
| GA at diagnosis (weeks) | 26.9 ± 4.2 |
| Estimated fetal weight discordance | 33.2% ± 7.9% |
| GA at delivery (weeks) | 32.3 ± 1.2 |
| Birth weight (g) |  |
| Normal cotwin | 1841.7 ± 368.3 |
| IUGR twin | 1213.3 ± 296.5 |
| Birth weight discordance | 34.2% ± 7.8% |
| A/REDV | 4 (66.7%) |
| Cesarean section | 6 (100.0%) |
| Female | 2 (33.3%) |
| NICU admission |  |
| Normal cotwin | 5 (83.3%) |
| IUGR twin | 6 (100.0%) |

*Note:* Discordance was calculated as (normal cotwin−IUGR twin)/normal cotwin × 100. Values represent n (%) or mean ± standard deviation. sIUGR, selective intrauterine growth restriction; IVF-ET, *in vitro* fertilization and embryo transfer; GA, gestational age; A/REDV, absent/reversed end diastolic velocity; NICU, neonatal intensive care unit.
