## Supplementary material for "The Proteome Landscape of Human Placentas for Monochorionic Twins with Selective Intrauterine Growth Restriction": Table S6.docx

| **Table S6 List of primers used for quantitative real-time PCR** | | |  |
| --- | --- | --- | --- |
| **Gene name** | **Gene description** | **Forward primer sequence (5'–3')** | **Reverse primer sequence (5'–3')** |
| *METAP2* | *methionyl aminopeptidase 2* | AAAGGACAAGAATGCGAATACCC | CAGGCTTGATCCAGCTCATTAC |
| *EFNB2* | *ephrin B2* | TATGCAGAACTGCGATTTCCAA | TGGGTATAGTACCAGTCCTTGTC |
| *VIM* | *vimentin* | GACGCCATCAACACCGAGTT | CTTTGTCGTTGGTTAGCTGGT |
| *MTDH* | *metadherin* | AAGCAGTGCAAAACAGTTCACG | GCACCTTATCACGTTTACGCT |
| *GAPDH* | *glyceraldehyde-3-phosphate dehydrogenase* | GGAGCGAGATCCCTCCAAAAT | GGCTGTTGTCATACTTCTCATGG |
