## Supplementary figures and images for "The Proteome Landscape of Human Placentas for Monochorionic Twins with Selective Intrauterine Growth Restriction"

### Figure S1.tif

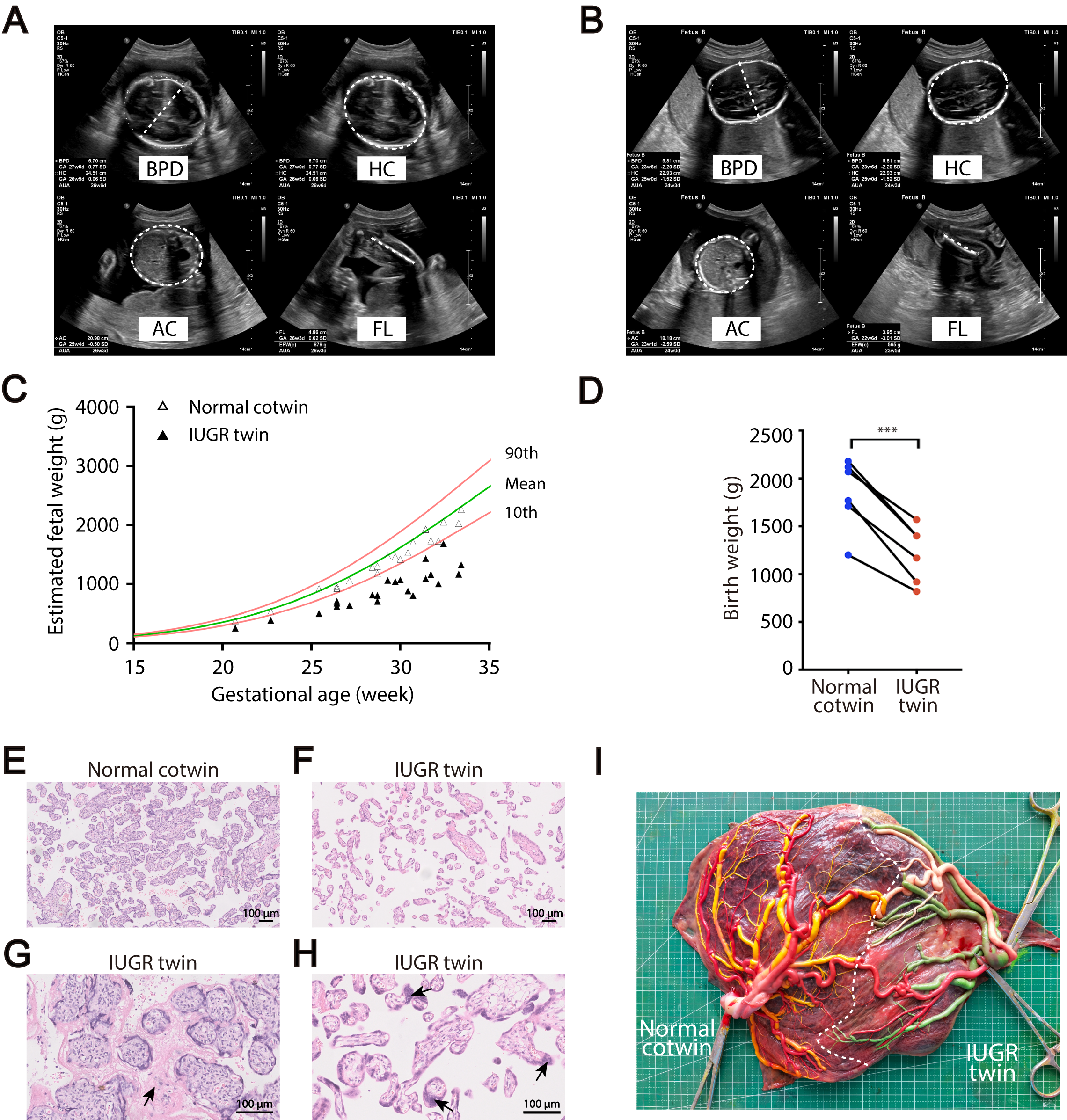

### Figure S2.tif

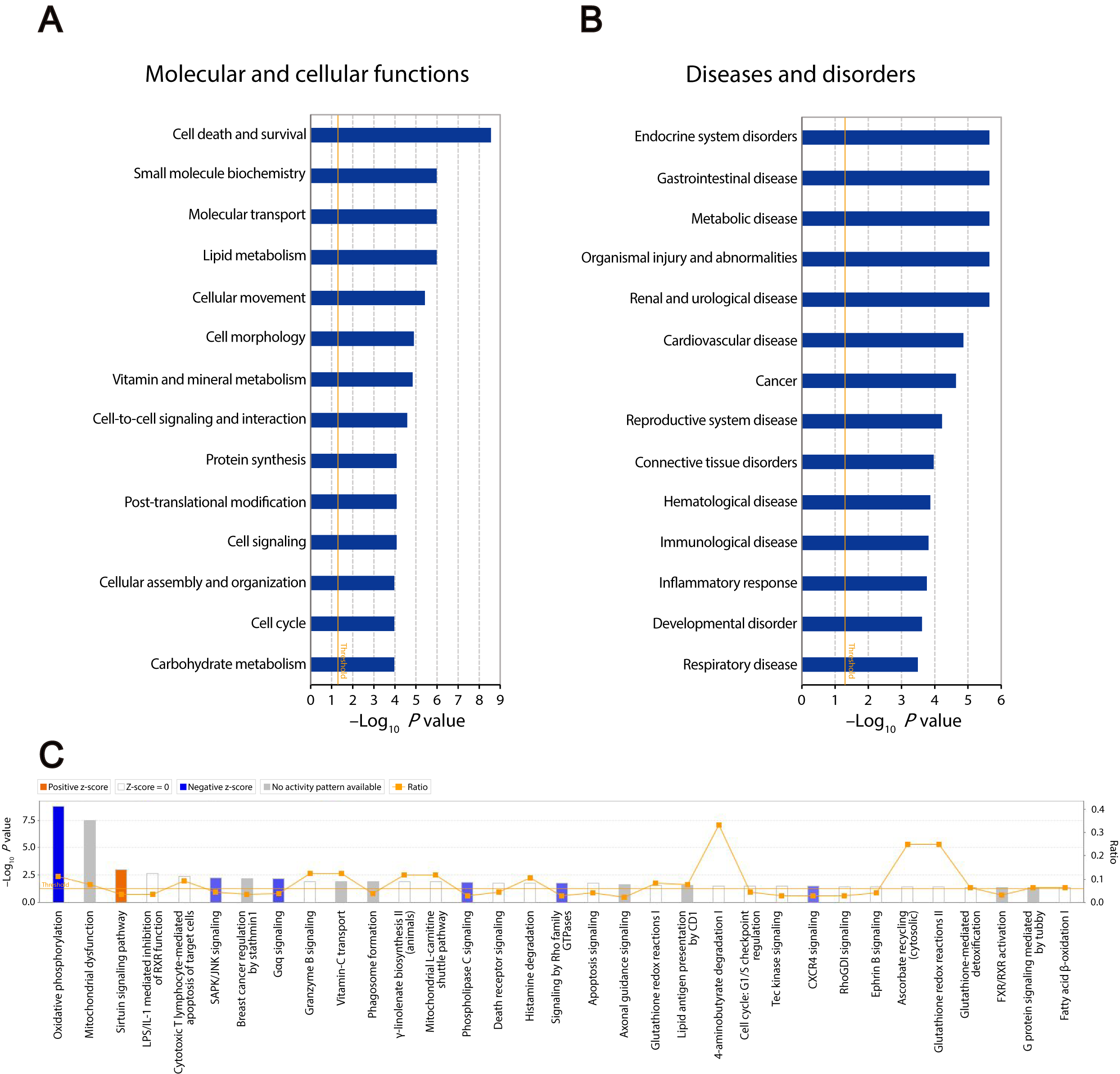
